# A relationship between Emotional intelligence and perceived parenting styles among undergraduate in Delhi University, India

**DOI:** 10.1101/2022.11.10.22282106

**Authors:** Dipti Mishra, Ranjana Singh

## Abstract

**Introduction:** Emotional Intelligence makes a major contribution and important factor in mental health. Parenting plays a pivotal role in training emotional intelligence because parents are responsible for the overall development of child. Not many studies have been done to find the association between emotional intelligence and parenting style. The objective of the study was to find the level of emotional intelligence in college students and if there is an association with perceived parenting style whether gender was as an effect modifier on the relationship between emotional intelligence and parenting style among young adults.

**Methods:** The study was conducted as a cross sectional study among college students in the age group of 18 to 24years. The study revealed that majority of the participants reported their parents parenting style as authoritative, followed by authoritarian.

**Result:** The proportion of authoritative father was 36% and authoritative mother was 37%. In this study the parenting style of mother was authoritative for both genders. The study results showed maternal authoritarian style (OR=0.3; 95%CI: 0.1, 0.5) and permissive style (OR=0.2; 95% CI: 0.2, 1.2) was negatively associated with high emotional quotient when compared to authoritative parenting style. Gender was an effect modifier in relationship between emotional intelligence and parenting style of father.

**Conclusion:** Emotional intelligence can be learned and modified and parents play a crucial rule in emotional development of child. They should acknowledge the developmental needs of child, listens to them, give them freedom and independence and also hold them accountable for their mistakes and behavior.

## Introduction

According to 2011 census in India, young people in the age group 15 to 29 years constitutes 27.53 percent of total population (1). Younger age group (12-24 years) is the beginning period for most of the mental disorders. Arnett calls the age group 18-25 as “emerging adulthood” (2). According to him, it is a period between adolescence and young adulthood. In this period many psychological changes are taking place.

Globally mental illnesses account for 30% non-fatal burden of disease and 10% of burden including disability and death. 20% of children and adolescents suffer from some form of mental disorder worldwide (3). In India, National Mental Health Survey estimated the current prevalence of mental illness in the age group of 18-29 years at 7.3% (excluding tobacco abuse) and lifetime prevalence of 9.54%(4). Emotional Intelligence is an important factor for positive mental health. It is defined in the literature as an ability to recognize, manage, distinguish, apply and understand emotions in a constructive way so is to relieve stress, understand with others, overcome challenges, communicate effectively, and eliminate conflict (5). Therefore, those Individuals who have developed capacity associated with emotional intelligence are able to perceive and convey their emotions, recognize the others emotions, utilize and regulate their temper and emotions to motivate flexible behavior(6).

Several studies have concluded that high emotional intelligence helps in positively focused coping mechanism and it is negatively associated with negatively focused coping strategy, low self-control, sensitivity, marital satisfaction, and cooperative responses (7)(8).Many studies have concluded that the emotional intelligence is a strong predictor of success in varied aspects such as mental wellbeing, physical health and academic achievements(9); (10); (11); (12).

There are many societal factors that influence emotional intelligence such as type of family, sex, family size, Birth order and parenting style. Parenting plays a pivotal role in training emotional intelligence because parents are responsible for the overall development of child. A parent helps in stimulating and nurturing the social, emotional, physical, and economical and intellect of the child from babyhood to maturity(13).

Researchers have found that a child wants a perfect balance between the level of affection and constraint they get from their parents in the home environment so as to influence social responsibility, high self-esteem, liberty, resiliency, proficiency, control and self-moving(14).

Depending upon the level of affection and constraint, there are three type of parenting style, namely, authoritative, authoritarian and permissive, as defined in the literature.

From the literature, it has been shown that parenting style plays an important role in emotional development of a child. Parenting style can be modified by effective intervention(15); (16).To the best of my knowledge there are hardly any studies in India which included college going young adults for assessing their emotional intelligence. When young adults come out from a school to a new college environment, dealing with social interaction, peer pressure and many other social activities becomes very crucial, in which having high emotional intelligence helps them. Hence the current study aims to assess their level of emotional intelligence and also to determine the role of gender as effect modifier on emotional intelligence and parenting style among young adults.

Also, the association between emotional intelligence and parenting style in majority of the available literature has been depicted using correlation analysis which holds a risk of confounded results as emotional intelligence is affected by other factors too(17); (18); (19). In the present study, the effect of parenting style on emotional intelligence was assessed after adjusting other factors also.

## Methods

### Study design and participants

A college based cross sectional survey was conducted among the undergraduate students. The study was conducted in the colleges of Delhi University. The study included 422 undergraduate college students (18 to 24 age) assuming prevalence of 53.2% of high level of emotional intelligence from previous literature. Students (both males and females) who were willing to participate and both parents alive were included in the study.

#### Sampling Method

A total of 6 colleges of south campus, DU were chosen based on the approval. Each college had three streams i.e. Science, arts and commerce. Two stage sampling methodology was adopted. Stage 1 where one undergraduate course from each stream was selected randomly and Stage 2 where students from these courses were conveniently selected. Total of 71 students was selected from each college.

### Measurement tools and methods

For collection of socio-demographic characteristics a questionnaire was designed by the researcher specifically for the study purpose. Pilot testing was done among 6 students to understand the feasibility of data collection and to understand if there are any issues in understanding the questions by participants. After piloting, appropriate modifications were done mainly in socio-demographic section. For socio-economic status Kupuswami scale was used.

For assessing emotional intelligence, the schutte emotional intelligence test was used. It consists of 33 items which are measured on five-point Likert scale where 1 stands for strongly disagree and 5 is for strongly agree, 3 items (numbers 5, 28 and 33) were reverse scored. Total score was calculated by adding scores of each question. Highest score was 165 and lowest score was 33. The score below 111 and score above 137 was considered as low emotional intelligence and high emotional intelligence, respectively(20). Assessment of parenting style was done by using parental authority question which was developed by Buri. It consists of 30 items, 10 questions for each parenting style and measured on 5-point Likert scale where 1 is for disagreeing strongly and 5 is for agreeing strongly. Total score for each parenting was calculated by adding the scores of each question. The parenting style with maximum score out of three was considered as the dominating parenting style of that parent.

### Statistical Analysis

Entry into excel was done on a regular basis during the data collection phase. After the entry was complete, data was then imported into Stata-15.1 version for Statistical analysis. Data cleaning and coding was done before analysis. The binary, nominal and ordinal variables are expressed in counts and percentages and the continuous variables are expressed along with Mean/Median and Standard deviation/Intra quartile range. The socio demographic profiles of the respondents are reported using descriptive statistics. Emotional intelligence was recorded as continuous variable initially but it was categorized into binary variable as high/moderate and low level of emotional intelligence to better reflect the proportion of emotional intelligence level. It was reported as proportion along with 95% confidence interval.

Parenting styles had three categories, namely authoritative, authoritarian and permissive, however, a fourth category was also there in the present study labeled as undefined parenting style where the score of two-parenting style was same. Sensitivity analysis was done to see if inclusion of this ‘undefined parenting style’ has any effect on the estimates. Since the estimates were not very different, it was decided to drop this category from the final analysis.

Apart from the key exposure variable, parenting style of father and mother, other variables used in the analysis were age, gender, occupation of father and mother, birth order and socio-economic status. Univariable and multivariable logistic regression was used to see association between parenting style and emotional intelligence after adjusting for the covariates. Interaction of gender was also investigated in the relationship of parenting and emotional intelligence and accordingly the final results are reported as odds ratio along with 95% Confidence Interval and p value.

## Results

### Socio demographic characteristics

The data for this study was collected from six colleges of Delhi University, South campus. The sample consists of 422 undergraduate students, the age ranges from 18 years to 24 years. The age was categorized into less than 19 years and more then equal to 19 years.

Tables 1 and 2 describes socio-demographic characteristics of participants along with family description and education and occupation of their parents.

**Table 1:** Demographic Profile of the participants

| **Participant's Characteristics** | **n=422** |
| --- | --- |
| **Age (%)** |  |
| **<19 Years** | 124(29.4) |
| **19 Years and above** | 298(70.6) |
| **Sex (%)** |  |
| **Male** | 222(52.6) |
| **Female** | 200(47.4) |
| **Religion (%)** |  |
| **Hindu** | 357(84.6) |
| **Non-Hindu** | 65(15.4) |
| **Stream (%)** |  |
| **Science** | 189(44.8) |
| **Arts** | 103(24.4) |
| **Commerce** | 130(30.8) |
| **Differential treatment of sibling by Gender (%)** |  |
| **Yes** | 55(13.0) |
| **Boarding school (%)** |  |
| Yes | 25(5.9) |

**Table 2:** Family description of participants and education and occupation of parents

| **Family Background of Participant's** | **N=422** |
| --- | --- |
| **Type of family (%)** |  |
| Nuclear | 300(71.1) |
| Joint | 122(28.9) |
| **Number of siblings (%)** |  |
| No sibling | 43(10.2) |
| One sibling | 239(56.6) |
| More than one siblings | 140(33.2) |
| **Birth order (%)** |  |
| First born | 171(40.5) |
| Middle born | 65(15.4) |
| Last born | 152(36.0) |
| Only child | 34(8.1) |
| **Marital status of parents (%)** |  |
| Married and living together | 400(94.8) |
| Married but living separately | 2(0.5) |
| Divorced and living alone | 2(0.5) |
| Single | 18(4.2) |
| **Head of the Family (%)** |  |
| Father | 375(88.9) |
| Mother | 22(5.2) |
| Grandfather | 22(5.2) |
| Grandmother | 2(0.5) |
| Uncle | 1(0.2) |
| **Socio-economic Status (%)** |  |
| **Upper class** | 181(42.9) |
| **Upper middle class** | 195(46.2) |
| **Lower middle class** | 43(10.2) |
| **Upper lower class** | 3(0.7) |
| **Education And Occupation of Parents** | N=422 |
| **Education of Mother (%)** |  |
| **No formal education** | 59(14.0) |
| **Primary level** | 60(14.2) |
| **Higher secondary** | 88(20.9) |
| **Graduation** | 125(29.6) |
| **Post-graduation and above** | 90(21.3) |
| **Occupation of Mother (%)** |  |
| **Non-professional** | 59(14) |
| **Professional** | 81(19.2) |
| **Home maker** | 282(66.8) |
| **Workplace of Mother (%)** |  |
| **In Delhi** | 367(87.0) |
| **Working outside Delhi** | 50(11.8) |
| **Working in Abroad** | 5(1.2) |
| **Education of father (%)** |  |
| **No formal education** | 0(0) |
| **Primary level** | 5(1.2) |
| **Higher secondary** | 60(14.2) |
| **Graduation** | 221(67.8) |
| **Post-graduation and above** | 136(32.2) |
| **Occupation of father (%)** |  |
| **Non-professional** | 122(28.9) |
| **Professional** | 298(69.2) |
| **Home maker** | 8(1.9) |
| **Workplace of Father (%)** |  |
| **In Delhi** | 321(76.1) |
| **Working outside Delhi** | 93(22.0) |
| **Working in Abroad** | 8(1.9) |

### Parenting styles of father and mother

The analyses showed that comparatively a large proportion of the participants have reported the parenting style of both their parents as authoritative. When parenting style of parents were looked separately among male and female, the analysis result showed that the perceived parenting style of father differ in both genders. Among males, 33% of the students perceived the parenting style of their father as authoritarian, where as in females, 42.5% of the students recognized their father’s style of parenting as authoritative. As far as mother’s parenting style was concerned, among females 39% of the students recognized their mothers’ style as authoritative, whereas 35% of male students recognized their mother’s parenting style as authoritative

### Emotional Intelligence

Highest score was 165 and lowest score was 33. The score below 111 and score above 137 was considered as low emotional quotient and high emotional quotient, respectively. The mean emotional intelligence level in entire sample was 120.1±16.0. The level of maximum emotional intelligence scored by subject was 157 and the level of minimum emotional intelligence scored by subject was 64.

Emotional intelligence was further categorized into Low EI (<111) and Moderate/high EI (>111), according to the Schutte emotional intelligence score.

From the study result it was found that 76% of the participants had moderately/high emotional quotient level and around 24% of students had low Emotional quotient. Figure 1 shows the distribution of emotional intelligence among males and females.

**Figure 1:**
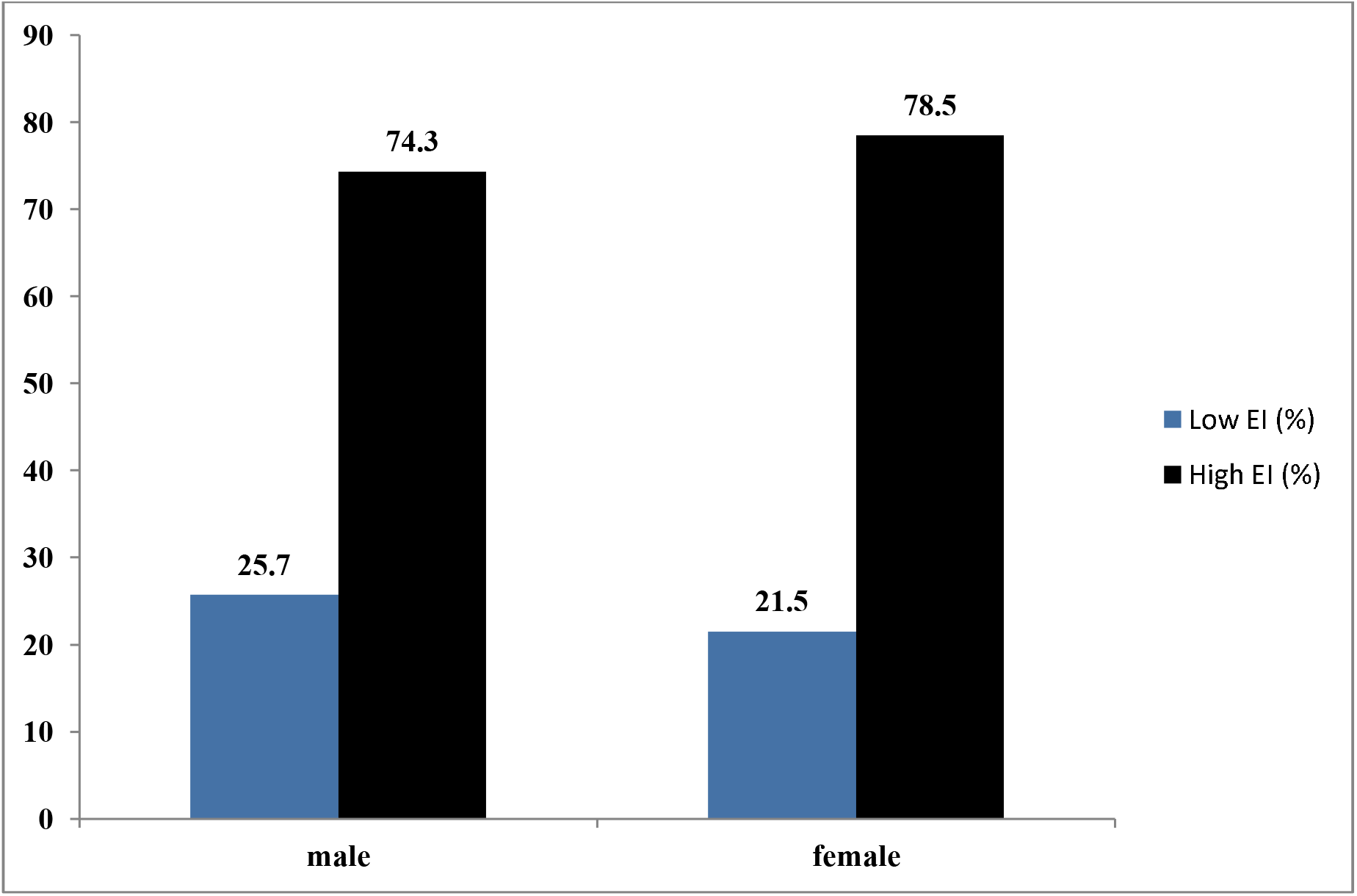
Emotional Intelligence among Males and Females

#### Distribution of emotional intelligence and parenting style of father and mother

Among males around 37% of those with high/moderate emotional quotient had authoritarian parenting style of father whereas among females, about 45% of those with high/moderate emotional quotient had authoritative parenting style of father. 40% of males had high/moderate emotional intelligence level in authoritative parenting style of mother and 39% of females had high/moderate level of emotional quotient in authoritative parenting style of mother.

### Emotional Intelligence and its associated Factors

Variables showing significant association with high/moderate level of emotional quotient in the univariable logistic regression are age and occupation of father. According to the results of univariable analysis, students of age 19 years and above have lower level of emotional quotient then student of 19 years. Higher level of emotional intelligence was seen in those students whose fathers have professional work when compared to those who are at home (Table 3).

**Table 3:**
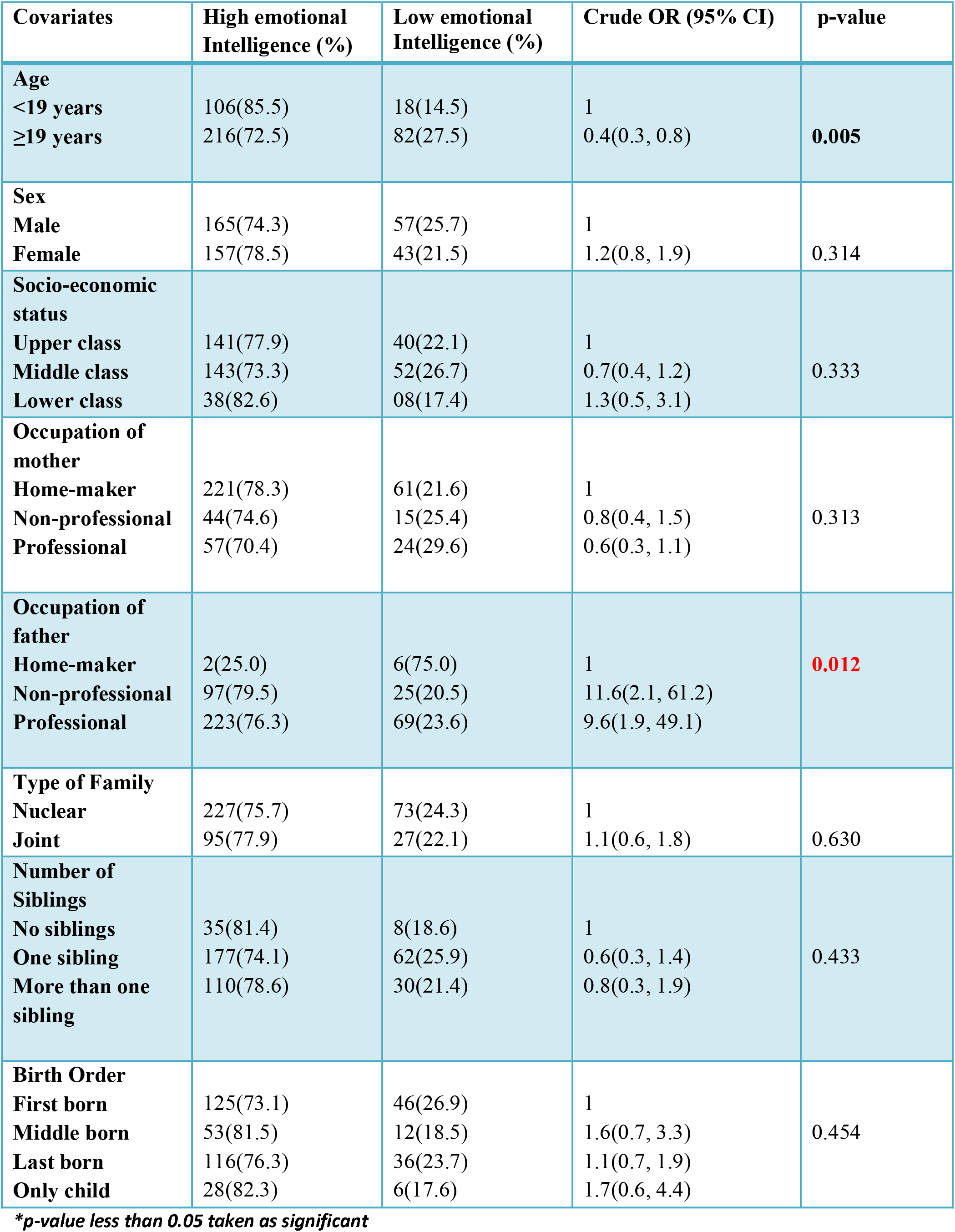
Univariable logistic regression analysis between emotional intelligence and other covariates (N=422)

#### Association between Emotional Intelligence and Parenting Style of Mother and Father- a multivariable Analysis

Emotional Intelligence was categorized into two categories Low EI (<111) and Moderate/high EI (>111), according to the Schutte emotional intelligence test. As the outcome variable EI was binary, multivariable logistic regression was performed for the adjusted association between parenting style and EI.

Parenting styles had three categories, namely authoritative, authoritarian and permissive, however, a fourth category was also there in the present study labeled as undefined parenting style where the score of two-parenting style was same. Sensitivity analysis was done to see if inclusion of this ‘undefined parenting style’ has any effect on the estimates. Since the estimates were not very different, it was decided to drop this category from the final analysis.

Apart from the key exposure variable, parenting style of father and mother, other variables used in the analysis were age, gender, occupation of father and mother, birth order and socio-economic status.

From the previous literature, it was found that parenting differs with gender which may influence the emotional intelligence of children also. Hence, interaction of gender with parenting style was investigated in the multivariable analysis separately for both the parents.

It was found that gender was an effect modifier between the relationship of emotional intelligence and parenting style of father (p value of interaction as 0.03) but not with the parenting style of mother. The Strata specific odds ratio for association between parenting style of father and emotional intelligence after adjusting for confounders was presented in Table 4. For association between emotional intelligence and parenting style of mother, gender was used as a confounder and the odds ratio was presented in Table 4.

**Table 4:** Multivariable logistic regression analysis for association between emotional intelligence and parenting style of father among male and female

| Parenting style of father | Male | Female |
| --- | --- | --- |
| Authoritative | 1 | 1 |
| Authoritarian | 2.1(0.9,5.1) | 0.7(0.3,1.8) |
| Permissive | 0.3(0.1,0.8) | 0.6(0.2,1.7) |

*\*Adjusted for age, occupation of father, socio-economic status and birth order*

From the analysis was, it was observed that males are more likely to have higher emotional intelligence when the perceived parenting style of father was authoritarian (OR=2.1; 95%CI: 0.9, 5.1) when compared to authoritative and permissive style of father, whereas the likelihood of high EI among females was less when the perceived parenting style of father was authoritarian or permissive when compared to authoritative parenting style of father.

The multivariable analysis between emotional quotient and parenting style of mother showed that in both permissive and authoritarian type of parenting style the odds of having high emotional intelligence was less in participants when compared to authoritative parenting style (Table 5).

**Table 5:** Multivariable logistic regression analysis for association between emotional Intelligence and parenting style of mother

| Parenting style of mother | Low Emotional intelligence<br>n(%) | Moderate/high Emotional Intelligence<br>n(%) | Crude OR (95% CI)<br>P-value | Adjusted OR (95% CI)<br>P-value |
| --- | --- | --- | --- | --- |
| Authoritative | 29(18.6) | 127(81.4) | 1 | 1 |
| Authoritarian | 24(23.8) | 77(76.2) | 0.5(0.3,0.8) | 0.3(0.1,0.5) |
| Permissive | 35(33.0) | 71(66.9) | 0.7(0.4,1.3) | 0.5(0.2,1.2) |
|  |  |  | **0.02\*** | **0.005\*** |

*p value <0.05 taken as significant Adjusted for age, gender, occupation of mother, birth order and socio-economic status*

## Discussion

There have been studies on emotional intelligence but not many studies to see association between perceived parenting style and emotional quotient among young adults. The main aim of this study was to assess the level of emotional intelligence and also to see association between emotional intelligence and perceived parenting style of mother and father. The sample comprised of 422 undergraduate college students in the age range of 18-24 years.

In the study, the mean score of emotional intelligence was 120 with standard deviation of 16. As reported by Schutte emotional intelligence scale, in large samples the mean score was around 124 with SD of 14 (20). This shows that mean of this study was almost equal to the original scale. The mean score obtained by this study was consistent with another study done in Australian college students of average age of 19 years(Emotional intelligence mean score = 123.8 with SD of 12.5) (9).

Another recent study conducted in Poland also reported mean score of emotional intelligence was 127.6 with standard deviation of 14.8 similar with the results of this study(21)

In this study emotional intelligence was categorized into binary variable for the purpose of better understanding and translation. Around 76% of participants had moderate/high level of emotional quotient, which was consistent with another study done among working young adults in Jorhat town(22). Another study was done in Haryana which showed that 57.08% participants had higher emotional intelligence and 42.91% participants had low emotional intelligence (23).

Previous research done on parenting style till now focuses on mainly three styles of parenting namely authoritative, permissive and authoritarian. In this study also these three categories of styles of parenting were evaluated using questionnaire on parental authority. Majority of the participants reported the parenting style of their parent’s as authoritative style, followed by authoritarian and small fraction reported it as permissive. The proportion of authoritative father was 36% and authoritative mother was 37%. The result from this study was consistent with previous study conducted among adolescents in Greece(24); (25). Another study conducted in Adolescents students in Haryana, India also showed that the majority respondents perceived their parents’ parenting style as authoritative followed by authoritarian style and permissive (23). Similar findings were also observed in another study done in Malaysia(26). Another study was done to see perception of parenting style in Indian students in college and U.S students in college. Majority of students in both counties perceived their own parents’ style as authoritative similar to what has been perceived in the present study(27).

In this study association between emotional intelligence and perceived parenting style of mother and father was analyzed separately. The result of this study revealed that the majority of the participants reported their father and mother parenting style as authoritative style, which was consistent with a study done in Karnataka on adolescents(17). Also, the result from the study revealed that permissive parenting style of both parents was negatively associated with emotional quotient, which was in accordance with the previous study results (28,29). This result was in accordance with the work of Alegre which found out the permissive parenting or low demand and high responsiveness was negatively related with the emotional intelligence(30) and authoritative style of parenting was positively related with emotional intelligence.

The parenting style differ across gender and its influence on emotional quotient. In this study the parenting style of mother was authoritative for both genders. The study results showed that the high level of emotional quotient was associated positively with authoritative style of parenting of mother in both genders, which was consistent with results found in previous study (16). Paternal parenting style differs across both genders in this study. Fathers are authoritative towards girls but authoritarian towards boys. Also, in boy’s paternal authoritarian style was significantly associated with high emotional quotient and in girl’s paternal authoritative style was positively related with high level of emotional intelligence. This finding corresponds to previous study where parents were authoritarian towards boys but authoritative towards girls (27). The same results were shown in the study done in Khasi adolescents where father was authoritative towards girls and strict towards boys(31) (17). The evidences from this study shows that parents are more warm towards female child than towards male child boys, while demonstrating more power over boys which was in accordance with pervious study (32).

There has been recent study done in Qazvin University of Medical Sciences in Qazvin, Iran, where it has been showed that emotional intelligence had positive association with conflict management strategies, and better verbal skills (33).

This study had little strength there were not many studies done in India to analyze the association between emotional intelligence and perceived parenting style among undergraduate students; hence this was one of its kinds. The response rate for this study was very 97%. A validated questionnaire was used to assess emotional intelligence and parenting style.

Although there were few limitations the questionnaire was a self-administered, hence that could have bled to bias from the participants side. Also, the questionnaire was lengthy which led participants responding quickly without paying attention and this may have affected the results. The colleges were chosen conveniently hence the generalization of the findings could be limited and as being cross-sectional study, it has all the limitations of a cross-sectional study design.

The following interpretations can be drawn from this study. There was a positive association between emotional quotient and perceived parenting style of parents. The dominating parenting style in both parents was authoritative style and more than 76% of participants had moderate/high emotional intelligence. Association between emotional intelligence and parenting style of mother showed that in both permissive and authoritarian type of parenting style the odds of having high emotional intelligence was less in participants when compared to authoritative parenting style, whereas for fathers parenting style, it was observed that males are more likely to have higher emotional intelligence when the perceived parenting style of father was authoritarian (OR=2.1; 95%CI: 0.9, 5.1), while the likelihood of high EI among females was more when the perceived parenting style of father was authoritative.

## Conclusion

As emotional intelligence is crucial for healthy living, therefore ways and means must be acknowledged to raise emotionally intelligent kid. Parents play a pivotal rule in emotional growth of child so parents should acknowledge the developmental needs of child, listens to them, give them freedom and independence and also hold them accountable for their mistakes and behavior.

## Data Availability

All data produced in the present study are available upon the reasonable request to he authors.

## Acknowledgement

I would like to express my deepest gratitude and acknowledgement to Indian Institute of Public Health and students of Delhi University for the completion of this study.

